# Hydrogen-rich jelly exhibits a positive effect on heart rate variability during orthostatic loading in healthy adults: A pilot randomized trial

**DOI:** 10.1101/2025.08.27.25334136

**Authors:** Yuki Muramoto, Genta Toshima, Emi Minaguchi, Kazuhisa Sugai, Kengo Nagashima, Yasunori Sato, Motoaki Sano, Kazuki Sato, Yoshinori Katsumata

**Affiliations:** Institute for Integrated Sports Medicine, Keio University School of Medicine, Tokyo, Japan; Clinical and Translational Research Center, Keio University Hospital, Tokyo, Japan; Department of Medicine and Clinical Science, Yamaguchi University Graduate School of Medicine, Yamaguchi, Japan

**Keywords:** Hydrogen-rich jelly, Orthostatic stress, Heart rate variability, Coefficient of variation of the R-R intervals, Low-frequency-to-high-frequency ratio, Autonomic recovery

## Abstract

Hydrogen-rich water has been shown to improve quality of life by enhancing mood, stress responsiveness, and autonomic regulation. The effects of hydrogen-rich jelly (HRJ) on autonomic responses during postural transition remain unclear. This double-blind, randomized, placebo-controlled crossover trial aimed to investigate the effects of HRJ on heart rate variability (HRV) indices in 50 healthy adults (21 males; mean age, 44.2 ± 11.3 years). HRJ (30–40 ppm, 10 g) was orally administered at 24 h, 1.5 h, and immediately before testing. The study protocol was approved by the Institutional Review Board of the Keio University School of Medicine (approval number: 20231194).HRV was continuously recorded during an orthostatic loading protocol consisting of four sequential 1-min phases: rest (seated baseline), stand-up (immediately after transitioning to standing), standing (sustained upright posture), and resitting (immediately after returning to sitting). We measured the coefficient of variation of the R–R interval (CVRR), low-frequency, high-frequency, and low-frequency-to-high-frequency (L/H) ratio components. Values for each posture were adjusted based on those at rest. Among the 48 participants with complete data under both conditions, HRJ intake resulted in a significantly lower CVRR during the stand-up phase (mean difference, −0.46; p = 0.047) and significantly lower L/H ratio during the resitting phase (mean difference, −1.51; p = 0.027). No significant differences were observed between the rest and standing phases. Our study suggests that HRJ intake modulates autonomic fluctuations during postural transitions.

## Introduction

Hydrogen gas suppresses chain-type cellular oxidative stress by selectively scavenging hydroxyl radicals [1], and its efficacy and safety in disease treatment and health maintenance are well established [2,3]. In the field of health promotion, hydrogen-rich water (HRW) attenuates oxidative stress and enhances parasympathetic activity, as reflected in heart rate variability (HRV) at rest [4]. Furthermore, HRW reduces psychological stress and improves HRV, both of which are characterized by decreased sympathetic nervous activity. Therefore, HRW may improve quality of life by influencing mood, stress responsiveness, and autonomic regulation [5,6]. With technological advancements, hydrogen can now be dissolved not only in water but also in jelly form, referred to as hydrogen-rich jelly (HRJ). A 10-g serving of HRJ provides an amount of hydrogen gas equivalent to 600 mL of HRW, enabling rapid intake of a substantial hydrogen dose in just a few seconds without the need to consume a large volume of liquid. However, the effects of HRJ intake on the autonomic nervous system (ANS) activity are not systematically investigated.

To date, evaluations of ANS balance have predominantly been conducted under resting conditions. In contrast, daily life involves dynamic fluctuations in physical activity across a spectrum of intensities that extend beyond periods of rest. Accordingly, the ability of the ANS to respond instantaneously and appropriately to these changes is essential for maintaining physiological stability during daily activities. Despite the importance of assessing reflexive autonomic responses to physical activity, research in this field remain limited [7]. A key barrier is the technical difficulty associated with assessing ANS activity during movement using conventional methods. HRV is typically analyzed using at least 5 min of resting electrocardiogram (ECG) data, employing a frequency domain analysis, such as the Fourier transform [8,9]. Therefore, evaluating HRV under dynamic conditions, where the heart rate exhibits rapid and transient fluctuations, poses a methodological challenge. A new system was developed to enable HRV analysis from just 30 s of ECG data using the maximum entropy method [8]. This approach has been validated through comparative studies using traditional 5-min HRV analysis protocols [9]. Building on this, a real-time HRV analysis system was developed to continuously evaluate HRV on a beat-to-beat basis [10-13], enabling the monitoring of HRV responses during representative physical activities in daily life, such as standing up and resitting [14,15].

Therefore, the present study aimed to investigate the effects of HRJ on ANS responses during mild everyday physical activities, specifically postural transitions such as standing up and resitting.

## Materials and methods

### Hydrogen gas content in the hydrogen jelly

The HRJ used in this study was manufactured in a facility certified under the Food Safety System Certification 22000 standard, which is a globally recognized food safety certification scheme approved by the Global Food Safety Initiative. The HRJ contained uniformly dispersed molecular hydrogen. Each portion was individually packaged in an aluminum pouch to preserve the hydrogen content. The concentration of dissolved hydrogen was measured using headspace gas chromatography and was confirmed to remain >30 ppm for up to 18 months after production (Supplemental Figure 1).

The ambient temperature for HRJ storage was maintained at 25 °C, ensuring a safe and stable environment. These conditions were sufficient to maintain the integrity of the hydrogen concentration in the stored jelly.

### Sample size

Based on preliminary analyses, the required sample size to detect changes in the HF component of HRV—an indicator of parasympathetic nervous system activity following the ingestion of HRJ—was estimated using G*Power. Assuming a two-tailed significance level of 0.05 and a statistical power of 0.90, the calculated sample size was 36 participants. However, the pilot data indicated that approximately 20% of the participants exhibited HF values exceeding three standard deviations above the mean, suggesting the presence of potential outliers. Therefore, the sample size was increased to 45 patients. Considering a projected dropout rate of 10% during the study period, 50 participants were included in the final recruitment.

### Participants

Fifty healthy adults with differing fitness levels, regardless of their exercise habits, were recruited between May and November 2024. The age of the participants ranged from 22 to 60 years, with a broad demographic range. The inclusion criteria were as follows: no underlying or pre-existing cardiovascular, respiratory, or metabolic diseases; no athletic injuries; no smoking habit; and no use of dietary supplements or medication of any type for at least 1 week prior to participation. The study protocol was approved by the Institutional Review Board of the Keio University School of Medicine (approval number: 20231194) and conducted in accordance with the principles of the Declaration of Helsinki. Written informed consent was obtained from all participants prior to study participation. This study was registered with the University Hospital Medical Information Network (UMIN000053818).

### Study outline

This was a double-blind, randomized, placebo-controlled trial with a crossover design. The study protocol consisted of three sessions. On the first day of participation (timing 1), body weight, body fat, and skeletal muscle mass were measured using a bioelectrical impedance analyzer (Inbody 470; Inbody Japan Inc., Tokyo, Japan). The participants were randomly assigned in a 1:1 ratio to receive either HRJ or PJ based on a randomization sequence generated in Microsoft Excel.

To maintain blinding, the jellies were provided in identical-looking pouches with a matching taste and appearance. The labels on the pouches were coded as ID_1 and ID_2, and the order of consumption was counterbalanced across the participants. Therefore, neither the participants nor the investigators could distinguish the intervention from the placebo. The participants consumed ID_1 prior to timing 1 and ID_2 prior to timing 2, with a washout period of at least 7 days between sessions to minimize carryover effects. The concentration of hydrogen in the body reaches its peak within a few minutes of hydrogen gas inhalation [16], and the hydrogen in exhaled breath is completely lost 60 min after intake [14]. Therefore, the participants were instructed to drink 10 g of HRJ or PJ within 2 min at the following time points: 24 h, 1.5 h, and immediately before the test. HRV was measured using a real-time HRV analysis system (CROSSWELL Corporation Co., Ltd., Kanagawa, Japan).

### Autonomic function test

HRV analysis during orthostatic stress was performed before the exercise load test. A wireless electrocardiograph (MWT-001, Arm Electronics, Tokyo) was used in the real-time HRV analysis system to continuously measure HRV under the following conditions:

1. Sitting (2 min): HRV was measured in a sitting posture.
2. Standing (2 min): After 2 min of sitting, the participants stood up as quickly as possible, and HRV was measured in a standing posture.
3. Resitting (1 min): After maintaining the standing posture, the participants sat back down, and HRV was measured in a sitting posture for 1 min.

HRV data from four time points were analyzed: rest, 1 min in the latter half of the sitting position; stand-up, 1 min immediately after standing; standing, 1 min in the latter half of the sitting position; stand-up, 1 min immediately after standing; standing, 1 min in the latter half of the standing position; and resitting, 1 min immediately after resitting (Supplemental Figure 2). Although the fast Fourier transform is commonly used for spectral analysis of R–R intervals, an accurate power spectral density from a short time-series fast Fourier transform is insufficient for estimating accurate power spectral densities from a short time series. Therefore, the MemCalc method was employed using HRV analysis software, which has been used in previous studies [14]. LF (0.05–0.15 Hz used as a mixed index of sympathetic and parasympathetic activities), HF (0.15–0.4 Hz, used as an index of parasympathetic activity), low-frequency-to-high-frequency (L/H), and CVRR components were used as HRV parameters [15].

In this study, ANS responses during each movement (stand-up, standing, and resitting) were evaluated in terms of recovery in the resting state. Thus, each autonomic parameter was expressed as a fluctuation in the resting value, representing the degree of deviation from the baseline condition.

### Study endpoints

The primary endpoint of this study was the HRV measured at 60 s in each postural phase at rest, stand-up, standing, and resitting. We compared the changes in the HRJ and PJ conditions and assessed this difference.

Additional subgroup analyses were performed to examine participant characteristics associated with favorable physiological responses to HRJ ingestion. Subgroups were defined based on the following nine characteristics: sex, smoking habits, alcohol consumption, physical activity habits, age, BMI, body fat percentage, and skeletal muscle mass. A favorable response was defined as an improvement in HRV parameters, particularly focusing on differences due to postural transitions from rest to resitting.

### Statistical analyses

Continuous variables are presented as means with standard deviations, and categorical variables are expressed as frequencies with percentages. HRV values exceeding three standard deviations from the mean were regarded as outliers and excluded from the analysis as missing values (with a maximum of one and a minimum of zero participants affected) [17].

We evaluated HRV between the HRJ and PJ conditions using mixed-effects models for repeated measures. Fixed effects included condition group, value at rest, timing, and interaction between condition group and timing. For outcomes at the rest phase, the value at rest was excluded from the fixed effects. An unstructured covariance matrix was assumed. If the model failed to converge, alternative covariance structures—heterogeneous first-order autoregressive, first-order autoregressive, and compound symmetry—were employed until convergence was confirmed. Differences between the HRJ and PJ conditions, along with 95% CIs, were estimated using least squares means. The Kenward–Roger method was used to estimate the variance of the parameter estimators and degrees of freedom.

Effect sizes were calculated using Cohen’s d, based on within-subject mean differences and standard deviations. Two-tailed p < 0.05 was considered statistically significant. Statistical analyses were performed using the SAS software (version 9.4, SAS Institute, Japan).

Fifty participants were enrolled in this study, with two withdrawing because of scheduling conflicts. The two conditions were assessed in a crossover design with a washout period of at least 7 (range, 7–14) days. Outliers exceeding three standard deviations were observed for some HRV parameters (range, 0–1 participant per variable). Therefore, statistical analyses were performed using data from 47–48 participants (Figure 1).

**Figure 1.**
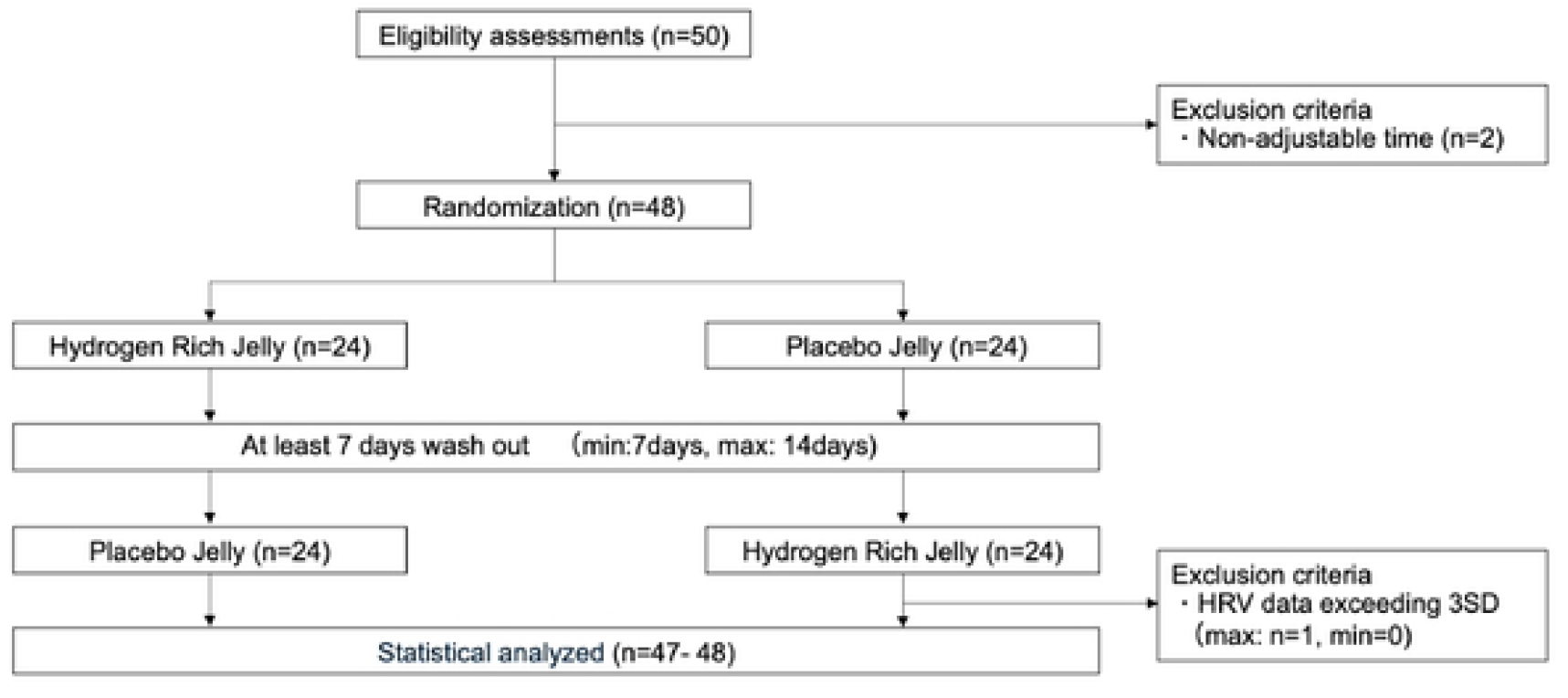
Consolidated Standards of Reporting Trials flow diagram of the study. Fifty participants were recruited for this study. Two were excluded owing to scheduling difficulties, resulting in 48 participants who were randomized in a crossover design to receive either hydrogen-rich or placebo jelly. All 48 participants completed an orthostatic loading protocol. The heart rate variability data were analyzed after excluding outliers that exceeded three standard deviations from the mean

## Results

The characteristics of the 48 participants retained for analysis were as follows: sex, 21 males (43.8%); mean age, 44.2 ± 11.3 years; and mean body mass index (BMI), 22.56 ± 3.69 kg/m^2^. Of these participants, 9 reported alcohol consumption (approximately 20 g/day) and 10 reported exercising more than four times per week (Table 1). No safety concerns regarding HRJ ingestion were identified throughout this study.

**Table 1.** Characteristics of all participants (N=48)

| Mean (SD) | All (n=48) |
| --- | --- |
| Age (years old) | 44.2 (11.3) |
| Height (cm) | 164.6 (7.9) |
| Mass (kg) | 61.28 (11.36) |
| BMI (kg/cm<sup>2</sup>) | 22.56 (3.69) |
| Body fat percentage (%) | 25.87 (8.78) |
| Lean body mass (kg) | 45.1 (8.51) |
| Skeletal muscle mass (kg) | 42.55 (8.06) |
| Muscle mass of right upper limb (kg) | 2.19 (0.64) |
| Muscle mass of left upper limb (kg) | 2.17 (0.61) |
| Muscle mass of trunk (kg) | 19.49 (3.82) |
| Muscle mass of right upper limb (kg) | 7.45 (1.57) |
| Muscle mass of right upper limb (kg) | 7.39 (1.52) |
| n (%) |  |
| Sex (Male) | 21 (43.8) |
| Habit of smoking | 3 (6.3) |
| Habit of drinking | 9 (18.8) |
| Habit of exercise(4times/week) | 10 (20.8) |

SD; standard deviation, BMI; body mass index

### Effects of HRJ on orthostatic stress responses adjusted for resting values

For the primary endpoint, during the stand-up phase, the coefficient of variation of the R–R interval (CVRR) was significantly lower in the HRJ condition compared with the placebo jelly (PJ) condition (mean difference = −0.458, 95% confidence interval [CI] = −0.910 to −0.006, p = 0.047). The low-frequency-to-high-frequency (L/H) ratio also tended to be lower in the HRJ condition than in the PJ condition (mean difference = −2.820, 95% CI = −6.012 to 0.372, p = 0.082), although this difference was not statistically significant (Table 2 and Figures 2 and 3). During the resitting phase, both the low-frequency (LF) component (mean difference = −74.549, 95% CI = −146.790 to −2.309, p = 0.043) and L/H ratio (mean difference = −1.506, 95% CI = −2.832 to −0.181, p = 0.027) were significantly lower in the HRJ condition than in the PJ condition (Table 2 and Figure 3 and 4). No significant differences in HRV parameters were observed between the conditions during the rest or standing phases.

**Table 2.** Effect of hydrogen water on orthostatic stress test heart rate variability analysis.

|  | Hydrogen jelly | Placebo |  |  |  |
| --- | --- | --- | --- | --- | --- |
| HR | Mean (SD) | Mean (SD) | Estimated mean difference (95%CI) | p-value | Cohen's d |
| Rest | 78.911 (13.143) | 79.743 (11.615) | -0.613 (-3.376, 2.149) | 0.657 | -0.07 |
| Stand-up | 6.707 (3.609) | 6.831 (3.538) | -0.093 (-0.935, 0.749) | 0.825 | -0.03 |
| Standing | 8.611 (4.552) | 8.713 (4.804) | -0.045 (-1.145, 1.055) | 0.934 | -0.02 |
| Resitting | 2.892 (2.723) | 3.111 (3.317) | -0.235 (-1.252, 0.783) | 0.644 | -0.07 |
| HF | Mean (SD) | Mean (SD) | Estimated mean difference (95%CI) | p-value | Cohen's d |
| Rest | 80.677 (58.486) | 86.325 (62.935) | 0.581 (-29.147, 30.309) | 0.969 | -0.09 |
| Stand-up | -26.41 (50.29) | -18.1 (48.642) | -4.991 (-19.922, 9.939) | 0.501 | -0.17 |
| Standing | -60.65 (64) | -60.7 (76.451) | -0.195 (-9.745, 9.354) | 0.967 | 0.00 |
| Resitting | -20.86 (61.98) | -14.64 (88.532) | -5.827 (-20.958, 9.303) | 0.442 | -0.08 |
| LF | Mean (SD) | Mean (SD) | Estimated mean difference (95%CI) | p-value | Cohen's d |
| Rest | 347.02 (270.49) | 437.48 (427.16) | 9.908 (-695.000, 714.820) | 0.888 | -0.26 |
| Stand-up | 44.566 (325.86) | 117.23 (326.55) | -50.518 (-135.520, 34.483) | 0.238 | -0.22 |
| Standing | -161.4 (372.53) | -163.5 (304.86) | 9.516 (-37.710, 56.742) | 0.687 | 0.01 |
| Resitting | -2.225 (433.55) | 102.41 (355.45) | -74.549 (-146.790, -2.309) | **0.043\*** | -0.26 |
| LH | Mean (SD) | Mean (SD) | Estimated mean difference (95%CI) | p-value | Cohen's d |
| Rest | 7.615 (10.268) | 7.874 (7.001) | 1.724 (-0.155, 3.603) | 0.071 | -0.03 |
| Stand-up | 2.591 (8.525) | 6.225 (11.261) | -2.820 (-6.012, 0.372) | 0.082 | -0.37 |
| Standing | 1.031 (6.115) | 3.131 (7.348) | -0.813 (-2.648, 1.022) | 0.377 | -0.31 |

| Resitting | 0.063 (5.99) | 2.975 (5.958) | -1.506 (-2.832, -0.181) | **0.027\*** | -0.49 |
| --- | --- | --- | --- | --- | --- |
| CVRR | Mean (SD) | Mean (SD) | Estimated mean difference (95%CI) | p-value | Cohen's d |
| Rest | 4.975 (1.563) | 5.178 (1.751) | 0.156 (-0.201, 0.513) | 0.384 | -0.12 |
| Stand-up | 1.073 (1.407) | 1.588 (1.996) | -0.458 (-0.910, -0.006) | **0.047\*** | -0.30 |
| Standing | -0.824 (1.044) | -0.643 (1.18) | -0.120 (-0.348, 0.107) | 0.293 | -0.16 |
| Resitting | 0.854 (1.567) | 1.23 (1.597) | -0.278 (-0.673, 0.118) | 0.164 | -0.24 |

\*, p < 0.05, 95%CI; 95 confidence intervals, SD; standard deviation, HR; Heart Rate,
HF = High Frequency, LF = Low Frequency, L/H =Low Frequency /High frequency, CVRR; Coefficient of Variation of R-R

**Figure 2.**
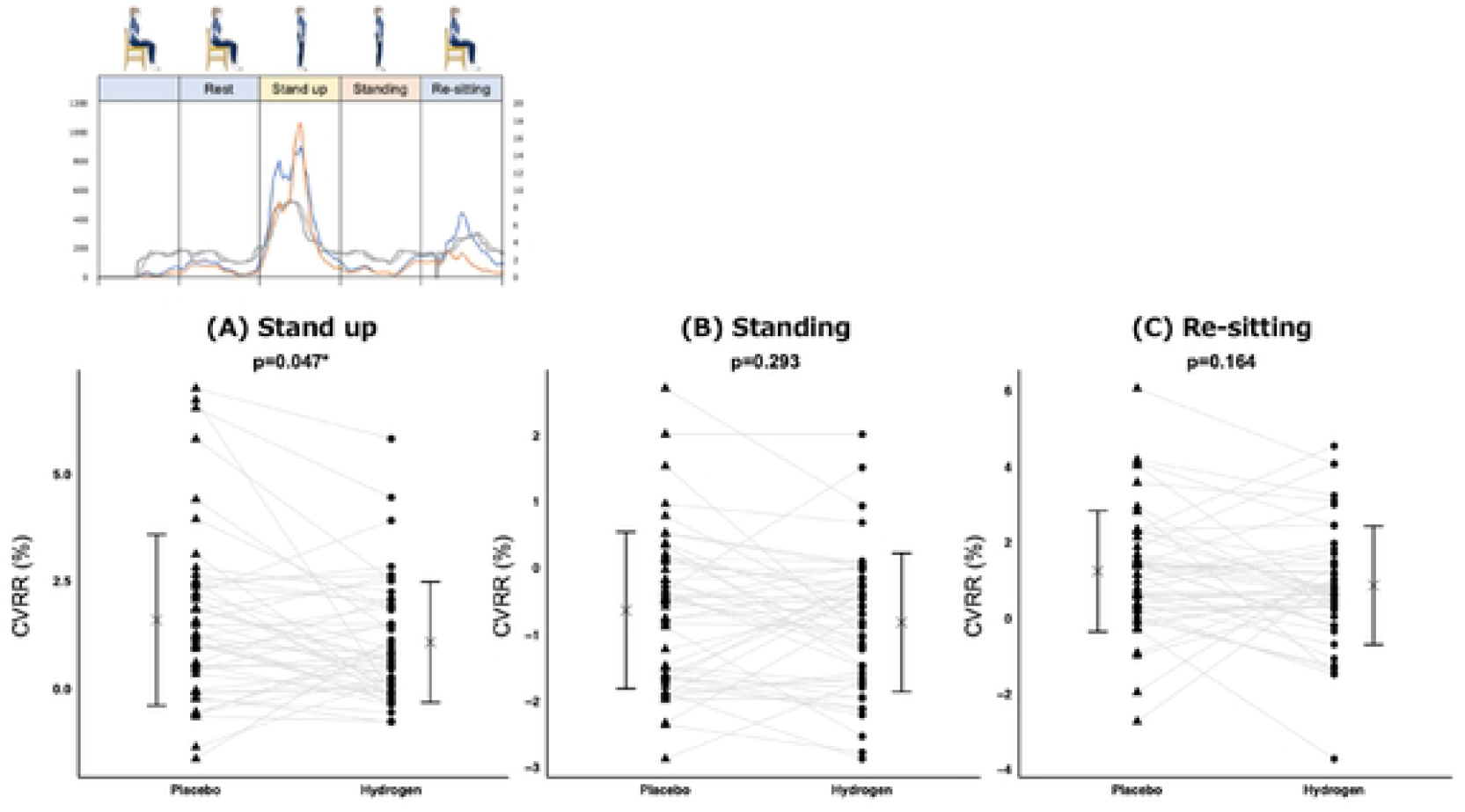
Coefficient of variation of the R–R interval (CVRR) results during the stand-up, standing, and resitting phases adjusted for rest. Mean ± standard deviation is displayed next to each plot to indicate the variability of the results. CVRR between the hydrogen-rich jelly (HRJ) and placebo jelly (PJ) conditions was evaluated using mixed-effects models for repeated measures. Panel A shows the results for the stand-up phase, panel B for the standing phase, and panel C for the resitting phase. CVRR was significantly lower under the HRJ condition than under the PJ condition during the stand-up phase. Illustration modified by the authors from free materials downloaded from ac-illust.com.

**Figure 3.**
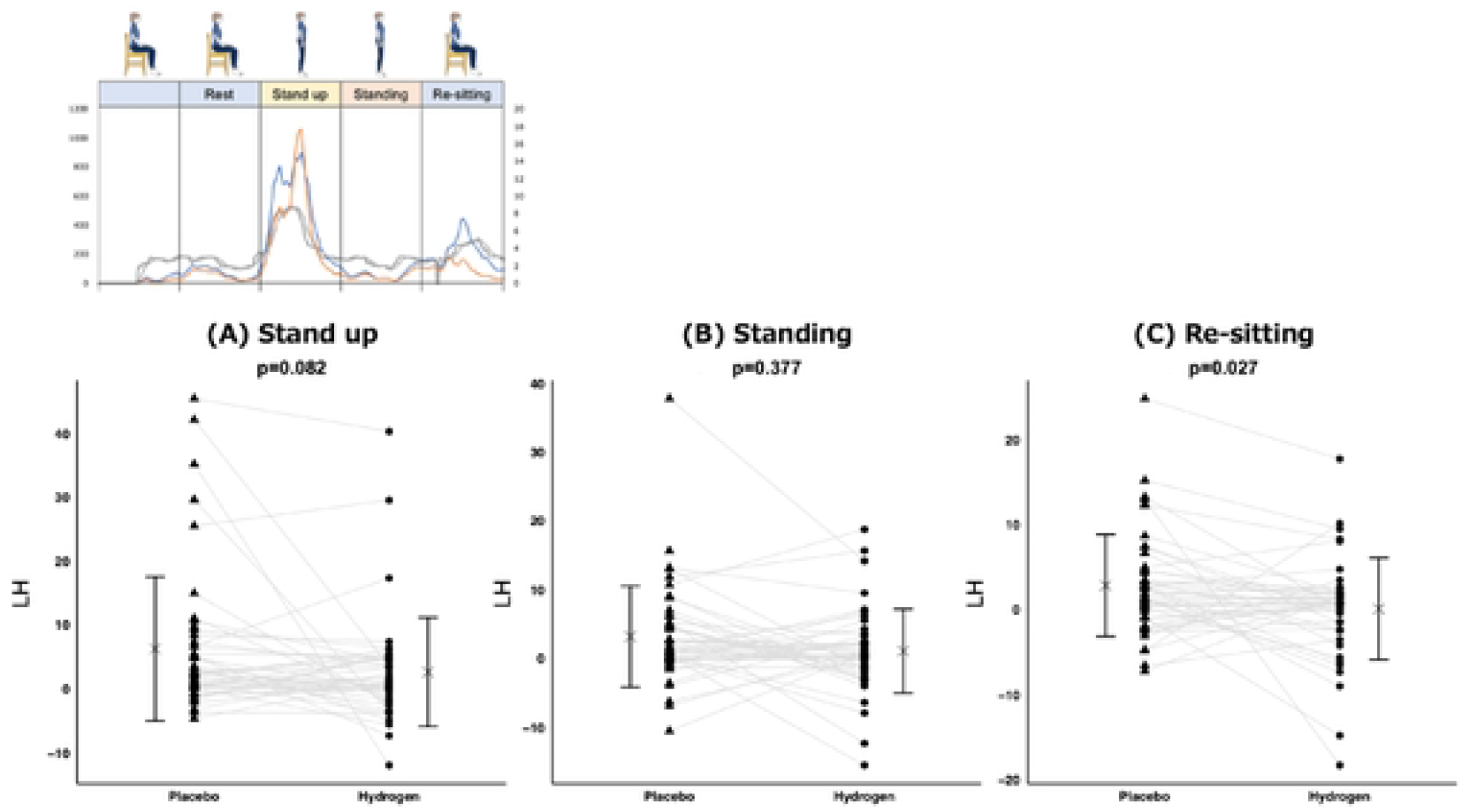
Low-frequency-to-high-frequency (L/H) ratio results during stand-up, standing, and resitting phases adjusted for Rest. Mean ± standard deviation is displayed next to each plot to indicate the variability of the results. The L/H ratio between hydrogen-rich jelly (HRJ) and placebo jelly (PJ) was evaluated using a mixed-effects model for repeated measures. Panel A shows the results for the stand-up phase, Panel B for the standing phase, and Panel C for the resitting phase. The L/H ratio was significantly lower under the HRJ condition than under the PJ condition during the resitting phase. Illustration modified by the authors from free materials downloaded from ac-illust.com.

**Figure 4.**
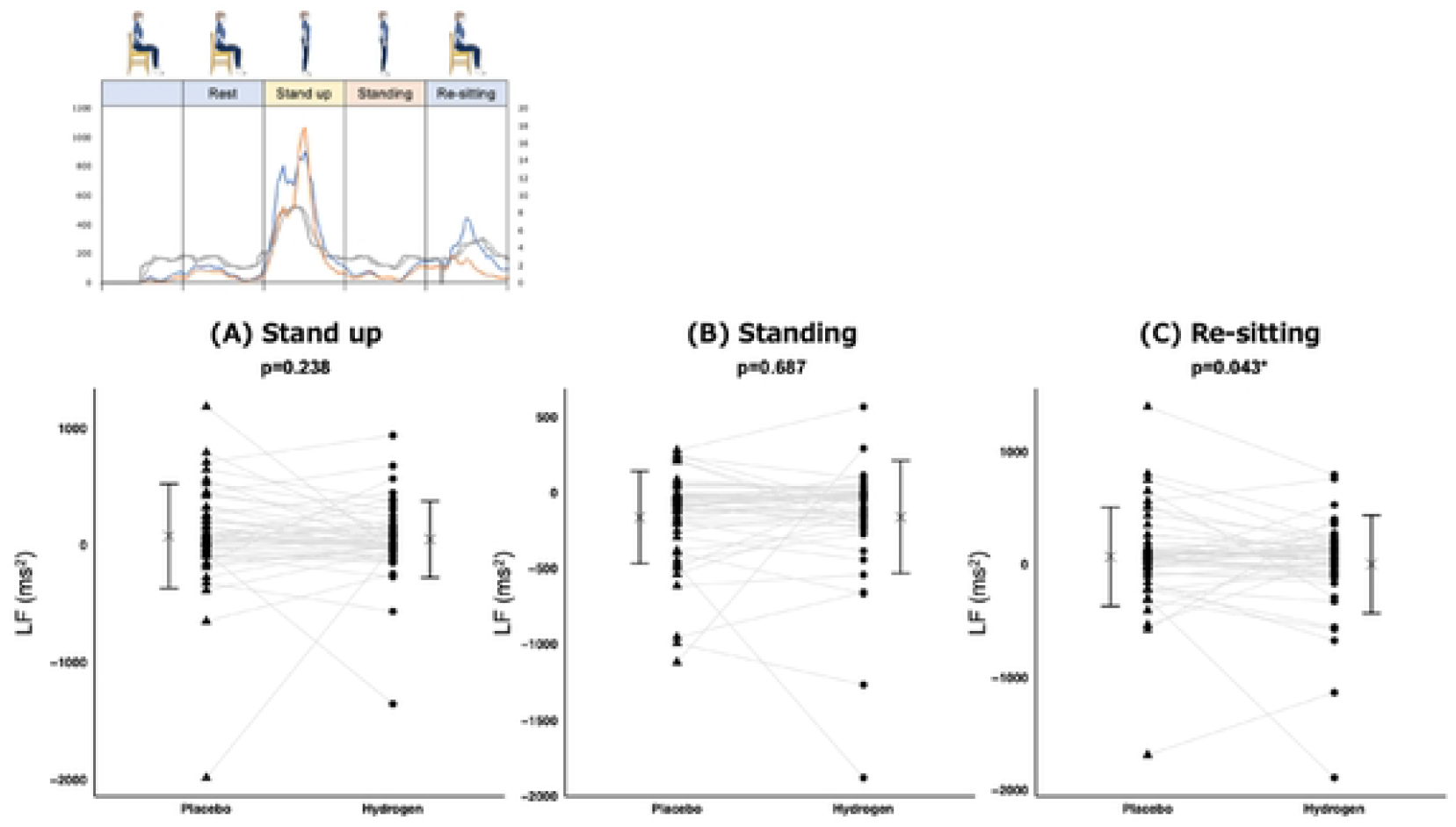
Low-frequency (LF) results during the stand-up, standing, and resitting phases adjusted for rest. Mean ± standard deviation is displayed next to each plot to indicate the variability of the results. The LF result between hydrogen-rich jelly (HRJ) and placebo jelly (PJ) was evaluated using mixed-effects models for repeated measures. Panel A shows the results for the stand-up phase, Panel B for the standing phase, and Panel C for the resitting phase. The LF value was significantly lower under the HRJ condition than under the PJ condition during the resitting phase. Illustration modified by the authors from free materials downloaded from ac-illust.com.

### Characteristics of participants showing improved heart rate variability following HRJ ingestion

Additional data analyses were performed to identify the subgroups of participants with reductions in CVRR and L/H ratio in the resitting phase following HRJ ingestion. For CVRR, significantly greater reductions were observed in female participants (mean difference = −0.525, 95% CI = −1.001 to −0.050) and in those with skeletal muscle mass below the median (mean difference = −0.598, 95% CI = −1.080 to −0.117) (Supplemental Figure 3). For L/H ratio, significant reductions were found among participants who consumed approximately 20 g of alcohol per day (mean difference = −2.044, 95% CI = −3.913 to −0.174), those younger than the median age (mean difference = −2.243, 95% CI = −3.623 to −0.863), and those with a BMI above the median (mean difference = −1.711, 95% CI = −3.259 to −0.163). Additionally, trends toward significance were observed for male participants (mean difference = −1.854, 95% CI = −3.828 to 0.121), those who exercised more than four times per week (mean difference = −2.060, 95% CI = −4.204 to 0.084), those with body fat percentages below the median (mean difference = −1.958, 95% CI = −4.183 to 0.704), and those with skeletal muscle mass above the median (mean difference = −1.686, 95% CI = −3.440 to 0.069) (Supplemental Figure 4).

## Discussion

This study revealed novel findings demonstrating that HRJ influences ANS responses during daily physical activities. During the stand-up phase (adjusted for resting values), the CVRR was significantly lower in the HRJ group than in the PJ group, with a trend toward lower L/H ratio. Furthermore, during the resitting phase (adjusted for resting values), both LF and L/H ratio components were significantly lower in the HRJ group than in the PJ group. No significant differences were observed between the resting and standing phases. These results suggest that HRJ intake alters the ANS response to orthostatic challenges, which supports the primary findings of the present study.

### Effects of HRJ on ANS changes between rest and orthostatic stress

CVRR is a key indicator of autonomic stability, and reduction in CVRR is associated with the progression of hypertension and diabetes [18]. CVRR recovery after exercise tends to be delayed in females, suggesting impaired parasympathetic reactivation during the post-exercise period [19]. These findings suggest that HRJ may enhance autonomic responsiveness during the stand-up phase.

The L/H ratio and LF component are indices associated with the sympathetic nervous system activity. Increases in these parameters during movement are considered normal physiological responses to exercise and stress. However, excessive elevations during physical activity may indicate heightened sympathetic activation or increased cardiovascular strain and are associated with autonomic imbalance, which is considered undesirable [20,21].

The ANS maintains a delicate balance influenced by both internal and external factors. Postural changes, such as standing up, induce immediate autonomic adjustments to maintain an upright posture [22,23]. Therefore, assessing both autonomic activity during stable states (i.e., rest and standing) and the rapid changes immediately following movement is crucial [24,25].

Accumulation of oxidative stress contributes to ANS imbalance [26-29]. Specifically, occupational stress and the intake of stimulants, such as caffeine, are associated with elevated L/H ratios and delayed recovery of HRV after physical activity [30,31]. Regarding the relationship between oxidative stress and the ANS, antioxidants containing high levels of vitamins promote HRV recovery [32]. Molecular hydrogen also exerts selective antioxidant effects by scavenging hydroxyl radicals [1,33]. Furthermore, HRW enhances the parasympathetic activity at rest and improves sleep quality in healthy individuals and young women [4-6]. In addition, a study in hypertensive rats demonstrated that inhalation of hydrogen gas for 1 h daily for 3 weeks led to decreased sympathetic activity and improved blood pressure control [34]. Our findings suggest that HRJ intake may contribute to improved recovery of sympathetic nervous system activity, potentially through its antioxidant effects.

However, we did not observe significant differences in the high-frequency (HF) component, a marker of parasympathetic nervous system activity. This finding contrasts with those of previous studies using HRW, which reported increased parasympathetic activity and decreased sympathetic activity following a 4-week intake of 600 mL/day HRW (0.8–1.2 ppm) [5]. In our study, the participants consumed molecular hydrogen at three separate time points: 24 h, 1.5 h, and immediately before the test—each time ingesting 10 g (30–40 ppm) of HRJ. These differing results may be attributable to the distinct intervention protocols, particularly the shorter duration and different patterns of hydrogen exposure, potentially leading to divergent effects on sympathetic and parasympathetic nervous system activities.

### Participant characteristics associated with improved ANS recovery following HRJ intake

The CVRR during the resitting phase, adjusted for resting values, was significantly lower in female participants who consumed HRJ. Notably, delayed CVRR recovery after exercise in females indicates reduced parasympathetic reactivation in this population [17]. The present findings suggest that HRJ intake may facilitate more efficient recovery of CVRR to resting levels in response to mild orthostatic challenges, such as standing up and resitting.

In contrast, among male participants, HRJ intake was associated with a more rapid return of L/H ratios to resting levels following the standing phase. This implies that HRJ may aid in the recovery of sympathetic nervous system activity after orthostatic challenge. Elevated L/H ratio is a risk factor for hypertension [35], and men tend to exhibit higher L/H ratios than women [36].

Considering these findings, HRJ may be particularly beneficial in supporting recovery from sympathetic overactivation in men. Notably, elevated L/H ratios have also been reported in men with habitual alcohol consumption [37], suggesting that HRJ may exert greater effects on this subgroup by promoting the return of L/H ratio to resting levels. That is, HRJ enhances reflexive autonomic recovery in a sex-specific manner, improving CVRR responses in female participants and L/H responses in male participants.

Moreover, physical activity habits and body composition, such as body fat percentage and skeletal muscle mass, are well-established factors that influence ANS function. Regular physical activity supports autonomic balance [38]. In contrast, excessive exercise is associated with reduced HRV and increased sympathetic activity, leading to autonomic imbalance [39]. However, the participants in this study were nonathletes and did not engage in exercise that would be considered excessive or likely to result in overtraining. Furthermore, increased body fat percentage and reduced skeletal muscle mass are associated with decreased parasympathetic activity and a shift toward sympathetic predominance, potentially disrupting autonomic balance [39,30]. Our findings suggest that HRJ may contribute to autonomic regulation by promoting L/H recovery, even in individuals with favorable autonomic profiles, such as those with moderate physical activity habits, lower body fat percentage, and relatively higher skeletal muscle mass.

## Limitations

This study has some limitations. First, HRV analyses of participants were performed under controlled conditions (at the same time of day and after the same meal) and with breathing rates standardized to 10–12 breaths per minute to minimize the confounding factors. Orthostatic loading was used to reduce the influence of the confounding factors. Second, this study did not consider the potential impact of the menstrual cycle on HRV in female participants, and information on menstrual status or the number of postmenopausal women was not collected. Given the potential impact of hormonal fluctuations during the menstrual cycle on ANS activity and HRV, future studies should address this limitation by collecting and analyzing these data.

Finally, we examined acute responses to HRJ intake. However, the effects of long-term HRJ intake on exercise and ANS function remain unknown and require further investigation.

## Conclusions

HRJ intake may facilitate faster recovery of sympathetic nervous system responses following movement. However, no significant changes in the HRV are observed at rest. Further studies are warranted to investigate the long-term effects of HRJ intake on ANS activity.

## Data Availability

All data from this study are presented in this manuscript or available from the corresponding author upon reasonable request.

## Acknowledgements

The authors would like to thank the participants who actively participated in this study and SOUKEN Corporation (https://www.souken-lab.co.jp/)for their help with recruitment. The manuscript was edited by Editage (www.editage.jp).

## Supplemental material

**Online Supplemental Figure 1. Hydrogen molecule content in hydrogen-rich jelly**

The concentration of dissolved hydrogen was measured using the headspace gas chromatography method and was confirmed to remain above 30 ppm for up to 18 months after production.

**Online Supplemental Figure 2. Heart rate variability (HRV) during the orthostatic loading test**

HRV data were analyzed at four time points: Rest – one minute during the latter half of the seated position; Stand up – the first minute immediately after standing; Standing – one minute during the latter half of the standing position; and Re-sitting – the first minute immediately after returning to the seated position. Illustration modified by the authors from free materials downloaded from ac-illust.com.

**Online Supplemental Figure 3. Participants showing lower CVRR during the re-sitting phase under the hydrogen-rich jelly (HRJ) condition**

The difference between the HRJ and PJ conditions and its 95% confidence interval was estimated using least squares means. The Kenward–Roger method was applied to estimate the variance of the parameter estimators and the degrees of freedom. A significantly lower CVRR under the HRJ condition was observed in female participants and those with skeletal muscle mass less than 40 kg.

**Online Supplemental Figure 4. Participants showing lower LH during the re-sitting phase under the hydrogen-rich jelly (HRJ) condition**

The difference between the HRJ and PJ conditions and its 95% confidence interval was estimated using least squares means. The Kenward–Roger method was used to estimate the variance of the parameter estimators and the degrees of freedom. A significantly lower LH under the HRJ condition was observed in participants who reported alcohol consumption of ≥20 g/day, were younger than 47 years, or had a BMI of ≥22.8 kg/m^2^. In addition, male participants, those who exercised ≥4 times per week, those with a body fat percentage <24.9%, and those with skeletal muscle mass ≥40 kg showed a tendency toward lower LH under the HRJ condition.

## References

1. Ohsawa, I. et al. Hydrogen acts as a therapeutic antioxidant by selectively reducing cytotoxic oxygen radicals. Nat. Med. 13, 688–694 (2007). doi:10.1038/nm1577

2. Sano, M. & Tamura, T. Hydrogen gas therapy: from preclinical studies to clinical trials. Curr. Pharm. Des. 27, 650–658 (2021). doi:10.2174/1381612826666201221150857

3. Katsumata, Y. et al. The effects of hydrogen gas inhalation on adverse left ventricular remodeling after percutaneous coronary intervention for ST-elevated myocardial infarction— first pilot study in humans. Circ. J. 81, 940–947 (2017). doi:10.1253/circj.CJ-17-0105

4. Botek, M., Sládečková, B., Krejčí, J., Pluháček, F. & Najmanová, E. Acute hydrogen-rich water ingestion stimulates cardiac autonomic activity in healthy females. Acta Gymnica 51, e2021–009 (2021). doi:10.5507/ag.2021.009

5. Mizuno, K. et al. Hydrogen-rich water for improvements of mood, anxiety, and autonomic nerve function in daily life. Med. Gas Res. 7, 247–255 (2017). doi:10.4103/2045-9912.222448

6. Fernández-Serrano, A. B. et al. Effects of hydrogen water and psychological treatment in a sample of women with panic disorder: a randomized and controlled clinical trial. Health Psychol. Res. 10, 35468 (2022). doi:10.52965/001c.35468

7. Swai, J., Hu, Z., Zhao, X., Rugambwa, T. & Ming, G. Heart rate and heart rate variability comparison between postural orthostatic tachycardia syndrome versus healthy participants; a systematic review and meta-analysis. BMC Cardiovasc. Disord. 19, 320 (2019). doi:10.1186/s12872-019-01298-y

8. Task Force of the European Society of Cardiology & North American Society of Pacing and Electrophysiology. Heart rate variability: standards of measurement, physiological interpretation and clinical use. Circulation 93, 1043–1065 (1996). doi:10.1161/01.CIR.93.5.1043

9. Wu, L., Shi, P., Yu, H. & Liu, Y. An optimization study of the ultra-short period for HRV analysis at rest and post-exercise. J. Electrocardiol. 63, 57–63 (2020). doi:10.1016/j.jelectrocard.2020.10.002

10. Okawara, H., Shiraishi, Y., Sato, K., Nakamura, M. & Katsumata, Y. Visually assessing work performance using a smartwatch via day-to-day fluctuations in heart rate variability. Digit. Health 10, 20552076241239240 (2024). doi:10.1177/20552076241239240

11. Shiraishi, Y. et al. Real-time analysis of the heart rate variability during incremental exercise for the detection of the ventilatory threshold. J. Am. Heart Assoc. 7, e006612 (2018). doi:10.1161/JAHA.117.006612

12. Ikura, H. et al. Real-time analysis of heart rate variability during aerobic exercise in patients with cardiovascular disease. Int. J. Cardiol. Heart Vasc. 43, 101147 (2022). doi:10.1016/j.ijcha.2022.101147

13. Ryuzaki, T. et al. Real-time estimation of anaerobic threshold during exercise using electrocardiogram in heart failure patients. J. Clin. Med. 12, 5225 (2023). doi:10.3390/jcm12165225

14. Sawada, Y. et al. New technique for time series analysis combining the maximum entropy method and non-linear least squares method: its value in heart rate variability analysis. Med. Biol. Eng. Comput. 35, 318–322 (1997). doi:10.1007/BF02534083

15. Shaffer, F., McCraty, R. & Zerr, C. L. A healthy heart is not a metronome: an integrative review of the heart’s anatomy and heart rate variability. Front. Psychol. 5, 1040 (2014). doi:10.3389/fpsyg.2014.01040

16. Mikami, T. et al. Drinking hydrogen water enhances endurance and relieves psychometric fatigue: a randomized, double-blind, placebo-controlled study. Can. J. Physiol. Pharmacol. 97, 857–862 (2019). doi:10.1139/cjpp-2019-0059

17. Wekenborg, M. K. et al. Determining the direction of prediction of the association between parasympathetic dysregulation and exhaustion symptoms. Sci. Rep. 12, 10648 (2022) doi: 10.1038/s41598-022-14743-4

18. Sammito, S. et al. Guideline for the application of heart rate and heart rate variability in occupational medicine and occupational health science. J. Occup. Med. Toxicol. 19, 15 (2024). doi:10.1186/s12995-024-00414-9

19. Viana, R. B. Sex differences in heart rate recovery and cardiac vagal reactivation following high-intensity functional training. J. Phys. Educ. Sport 24, 251 (2024). doi:10.7752/jpes.2024.09251

20. Kim, HH. G., Cheon, E. J., Bai, D. S., Lee, Y. H. & Koo, B. H. Stress and heart rate variability: a meta-analysis and review of the literature. Psychiatry Investig. 15, 235–245 (2018). doi:10.30773/pi.2017.08.17

21. Akizuki, H. & Hashiguchi, N. Heart rate variability in patients presenting with neurally mediated syncope in an emergency department. Am. J. Emerg. Med. 38, 211–216 (2020). doi:10.1016/j.ajem.2019.02.005

22. Schiweck, C., Piette, D., Berckmans, D., Claes, S. & Vrieze, E. Heart rate and high frequency heart rate variability during stress as biomarker for clinical depression: a systematic review. Psychol. Med. 49, 200–211 (2019). doi:10.1017/S0033291718001988

23. Okamura, S. et al. Peripheral arterial tone during active standing. Pflugers Arch. 473, 1939–1946 (2021). doi:10.1007/s00424-021-02632-0

24. Fukuda, S. et al. A potential biomarker for fatigue: oxidative stress and anti-oxidative activity. Biol. Psychol. 118, 88–93 (2016). doi:10.1016/j.biopsycho.2016.05.005

25. Ishihara, I. et al. Effect of work conditions and work environments on the formation of 8-OH-dG in nurses and non-nurse female workers. J. UOEH 30, 293–308 (2008). doi:10.7888/juoeh.30.293

26. Lee, J.-S., Kim, H.-G., Lee, D.-S. & Son, C.-G. Oxidative stress is a convincing contributor to idiopathic chronic fatigue. Sci. Rep. 8, 12890 (2018). doi:10.1038/s41598-018-31270-3

27. Yang, S. & Lian, G. ROS and diseases: role in metabolism and energy supply. Mol. Cell. Biochem. 467, 1–12 (2020). doi:10.1007/s11010-019-03667-9

28. Borchini, R. et al. Heart rate variability frequency domain alterations among healthy nurses exposed to prolonged work stress. Int. J. Environ. Res. Public Health 15, 113 (2018). doi:10.3390/ijerph15010113

29. Benjamim, C. J. R. et al. Caffeine slows heart rate autonomic recovery following strength exercise in healthy subjects. Rev. Port. Cardiol. (Engl. Ed.) 40, 399–406 (2021). doi:10.1016/j.repc.2020.07.021

30. Bellafiore, M. et al. Redox and autonomic responses to acute exercise-post recovery following Opuntia ficus-indica juice intake in physically active women. J. Int. Soc. Sports Nutr. 18, 43 (2021). doi:10.1186/s12970-021-00444-2

31. Iketani, M. & Ohsawa, I. Molecular hydrogen as a neuroprotective agent. Curr. Neuropharmacol. 15, 324–331 (2017). doi:10.2174/1570159x14666160607205417

32. Sugai, K. et al. Daily inhalation of hydrogen gas has a blood pressure-lowering effect in a rat model of hypertension. Sci. Rep. 10, 20173 (2020). doi:10.1038/s41598-020-77349-8

33. Fagard, R. H. Exercise characteristics and the blood pressure response to dynamic physical training. J. Hypertens. 19, 389–397 (2001). doi:10.1097/00004872-200103000-00006

34. Yugar, L. B. T. et al. The role of heart rate variability (HRV) in different hypertensive syndromes. Diagnostics (Basel) 13, 785 (2023). doi:10.3390/diagnostics13040785

35. Mori, H. et al. Heart rate variability and blood pressure among Japanese men and women: a community-based cross-sectional study. Hypertens. Res. 37, 779–784 (2014). doi:10.1038/hr.2014.73

36. Addleman, J. S., Lackey, N. S., DeBlauw, J. A. & Hajduczok, A. G. Heart Rate Variability Applications in Strength and Conditioning: A Narrative Review. J. Funct. Morphol. Kinesiol. 9, 93 (2024). doi:10.3390/jfmk9020093

37. Mourot, L. et al. Decrease in heart rate variability with overtraining: assessment by the Poincaré plot analysis. Clin. Physiol. Funct. Imaging 24, 10–18 (2004). doi:10.1046/j.1475-0961.2003.00523.x

38. Cvijetic, S., Macan, J., Boschiero, D. & Ilich, J. Z. Body fat and muscle in relation to heart rate variability in young-to-middle age men: a cross sectional study. Ann. Hum. Biol. 50, 108–116 (2023). doi:10.1080/03014460.2023.2180089.

39. Navarro-Lomas, G., Plaza-Florido, A., De-la-O, A., Castillo, M. J. & Amaro-Gahete, F. J. Genetic and environmental determinants of autonomic nervous function in adolescents. Am. J. Hum. Biol. 35, e23945 (2023). doi:10.1002/ajhb.23945

40. Hirano, M. et al. Pharmacokinetics of hydrogen administered intraperitoneally as hydrogen-rich saline and its effect on ischemic neuronal cell death in the brain in gerbils. PLOS One 17, e0279410 (2022). doi:10.1371/journal.pone.0279410

